# Sociodemographic and health-related differences in undiagnosed hypertension in the Health Survey for England 2015-2019

**DOI:** 10.1101/2023.07.03.23292175

**Authors:** Emma Campbell, Ellie Macey, Chris Shine, Vahé Nafilyan, Nathan Cadogan Clark, Piotr Pawelek, Isobel Ward, Andrew Hughes, Veena Raleigh, Amitava Banerjee, Katie Finning

## Abstract

**Background:** Hypertension is a leading cause of morbidity and mortality worldwide, yet a substantial proportion of cases are undiagnosed. Understanding the scale of undiagnosed hypertension and identifying groups most at risk is important to inform approaches to detection.

**Methods:** We used data from the 2015 to 2019 Health Survey for England, an annual, cross-sectional, nationally representative survey. Undiagnosed hypertension was defined by a measured blood pressure of 140/90mmHg or above but no history of diagnosis. Age-adjusted prevalence of undiagnosed hypertension was estimated across sociodemographic and health-related characteristics. To assess the independent association between undiagnosed hypertension and each characteristic, we fitted a logistic regression model adjusted for sociodemographic factors.

**Findings:** An estimated 30.7% (29.0-32.4%) of men and 27.6% (26.1-29.1%) of women with hypertension were undiagnosed. Younger age, lower Body Mass Index, and better self-reported general health were associated with an increased likelihood of hypertension being undiagnosed for men and women. Living in rural areas and in regions outside of London and the East of England were also associated with an increased likelihood of hypertension being undiagnosed for men, as were being married or in a civil partnership and having higher educational qualifications for women.

**Interpretation:** Hypertension is commonly undiagnosed, and some of the groups less at risk of hypertension are most likely to be undiagnosed. Given the high lifetime risk of hypertension and its strong links with morbidity and mortality, our findings suggest a need for greater awareness of the potential for undiagnosed hypertension, including among those typically considered ‘low risk’.

## Introduction

Hypertension is a leading cause of morbidity and mortality worldwide and is responsible for between 8 and 10 million deaths each year. ^[1–3]^ High blood pressure accounted for an estimated 9% of global Disability-Adjusted Life Years in 2019. ^[4]^ In England, 28% of adults had hypertension in 2019, defined as having a measured blood pressure of 140/90 mmHg or above or self-reporting antihypertensive medication use. ^[5]^ Hypertension also places considerable burden on health services and is responsible for 12% of all visits to General Practitioners in England, with an estimated annual cost to the National Health Service of over £2 billion. ^[6]^

Lowering blood pressure is a well-established strategy for reducing the incidence of cardiovascular diseases such as stroke, myocardial infarction, and coronary heart disease, and is associated with a significant reduction in all-cause mortality. ^[7–8]^ Early identification and treatment are associated with significant health and economic benefits because they reduce the need for more elaborate and costly interventions required to treat the complications of hypertension. ^[2–3]^ Hypertension prevention focuses on lifestyle interventions such as eating a healthy diet, taking regular exercise, and reducing alcohol consumption, and the National Institute for Health and Care Excellence (NICE) in England recommends the same lifestyle interventions for individuals diagnosed with hypertension. ^[9]^ This is followed by antihypertensive medication if blood pressure is not successfully controlled through lifestyle interventions alone.

Hypertension rarely causes symptoms in the early stages and a substantial proportion of cases are undiagnosed. ^[2–3]^ A review of hypertension trends between 1990 and 2019 found that 41% of women and 51% of men with hypertension globally did not report a diagnosis, ^[2]^ and previous estimates suggest that around one third of hypertension cases in England are undiagnosed. ^[10]^ In 2019, a new coalition led by Public Health England and NHS England outlined plans to improve detection and prevention of the major causes of cardiovascular disease, including an ambition to diagnose 80% of hypertension cases by 2029. ^[11]^ However, there was a substantial decrease in the proportion of hypertension cases diagnosed and treated during the COVID-19 pandemic, leading to a backlog of care and an urgent need to identify those who are undiagnosed. ^[12–13]^

Previous research has identified groups who are at increased risk of hypertension. This includes older adults, males, individuals belonging to Black ethnic groups, and those living in more deprived areas, as well as those with known health-related risk factors such as being overweight or physically inactive. ^[3, 5, 14–15]^ However, less is known about which groups are most likely to be undiagnosed. Better understanding of inequalities in undiagnosed hypertension could lead to improved rates of diagnosis through targeted screening or public health campaigns. This study aims to identify sociodemographic and health-related differences in undiagnosed hypertension using a nationally representative sample of adults in England.

## Methods

### Study population

We used data from the Health Survey for England (HSE), an annual cross-sectional survey of people living in private households in England. ^[5]^ We pooled data from the 2015 to 2019 surveys, which were the most recent data years available at the time of analysis. The HSE follows a multi-stage stratified probability sampling design; full details of the sampling methods are available via NHS Digital. ^[5]^ All participants complete a health interview, which includes questions on demographic characteristics and health history. Participants are then visited by a study nurse who takes measurements such as blood pressure. Prior to 2018 all participants were eligible for a nurse visit and in 2018-2019 89% of surveyed addresses were randomly selected for a nurse visit. However, some eligible participants did not complete the nurse visit. The proportion of adult participants who completed a nurse visit between 2015-2019 ranged from 59% to 67%. Our analytical sample was restricted to respondents from the 2015-2019 HSE who were aged 16 years and over, were not pregnant at the time of interview, and had valid blood pressure data. Supplementary Material Figure S1 shows the flow of survey respondents.

### Outcomes

#### Self-reported diagnosis of hypertension

Participants were asked “Do you now have, or have you ever had high blood pressure (sometimes called hypertension)?”. If answered yes, participants were asked “Were you told by a doctor or nurse that you had high blood pressure?”. We classified participants as having self-reported diagnosed hypertension if they answered ‘yes’ to both questions and the high blood pressure didn’t occur only during pregnancy.

#### Blood pressure measurement

During the nurse visit, three consecutive blood pressure readings were taken. We used the mean of the second and third measurements, in line with previous research and guidelines. ^[10, 16]^ We classified participants as having high blood pressure if their systolic pressure was 140 mmHg or above or their diastolic pressure was 90 mmHg or above.

#### Total hypertension and undiagnosed hypertension

We used the variables above to derive our two outcomes of interest:

a. Total hypertension - individuals who self-reported a diagnosis of hypertension or had a blood pressure measurement of 140/90 mmHg or above
b. Undiagnosed hypertension – individuals whose blood pressure measurement was 140/90 mmHg or above but who did not self-report a diagnosis of hypertension

### Predictors

We explored a variety of sociodemographic and health-related predictors of hypertension and undiagnosed hypertension, based on the literature, theoretical relevance, and data availability. These were: sex, age group, ethnicity, region, rural-urban classification, relationship status, highest educational qualification, National Statistics Socio-Economic Classification (NS-SEC), Body Mass Index (BMI), self-reported general health, and smoking status. These were all collected during the health interview. Details of data collection methods and variable levels for all predictors are available in Supplementary Material Table S1.

### Statistical analysis

All analyses accounted for the survey design and incorporated weights that accounted for selection, non-response, and population profile in the main sample, as well as non-response bias introduced through the nurse visits. ^[5]^ Weights were re-scaled to the pooled dataset and sub-sample of interest (adults who were not pregnant and had valid blood pressure data), using guidance provided by the HSE survey team. For each predictor we estimated the age-adjusted prevalence of (a) total hypertension among all adults and (b) undiagnosed hypertension among those with hypertension, by fitting logistic regression models adjusted for age (in five-year age bands, the most granular level available). Marginal means were then used to estimate the age-adjusted prevalence with hypertension and undiagnosed hypertension in each group. Second, we examined the odds of hypertension being undiagnosed for each of our predictors after adjusting for other factors. We fitted minimally adjusted logistic regression models (adjusted for age and stratified by sex) followed by fully adjusted models that adjusted for age, ethnicity, region, urban-rural classification, relationship status, highest educational qualification, and NS-SEC. Statistically significant interactions were identified between sex and several of our predictor variables, therefore all analyses were stratified by sex. Analyses were conducted using R 4.0.2.

### Ethics

Ethical approval was granted by the UK Statistics Authority Data Ethics Team, falling under the National Statistician’s Data Ethics Advisory Committee.

## Results

### Characteristics of the sample

The sample involved 21,476 individuals; 55.8% were female, 89.3% reported a White ethnic background. Table 1 summarises the characteristics of the sample. There was less than 0.2% missing data on all variables except BMI, which was missing for 2,149 individuals (10% of the sample).

**Table 1.** Characteristics of the sample

|  |  | **Sample N (%)** |
| --- | --- | --- |
| **Sex** | Male | 9,485 (44.2) |
|  | Female | 11,991 (55.8) |
| **Age group** | 16-24 years | 1,438 (6.7) |
|  | 25-34 years | 2,599 (12.1) |
|  | 35-44 years | 3,268 (15.2) |
|  | 45-54 years | 3,643 (17.0) |
|  | 55-64 years | 3,776 (17.6) |
|  | 65-74 years | 3,894 (18.1) |
|  | 75 years and over | 2,858 (13.3) |
| **Ethnicity** | White | 19,156 (89.3) |
|  | Black | 527 (2.5) |
|  | Asian | 1,340 (6.2) |
|  | Other | 438 (2.0) |
| **Region** | East Midlands | 2,015 (9.4) |
|  | East of England | 2,586 (12.0) |
|  | London | 2,262 (10.5) |
|  | North East | 1,881 (8.8) |
|  | North West | 2,826 (13.2) |
|  | South East | 3,618 (16.8) |
|  | South West | 2,281 (10.6) |
|  | West Midlands | 2,055 (9.6) |
|  | Yorkshire and the Humber | 1,952 (9.1) |
| **Rural-urban classification** | Urban | 16,884 (78.6) |
|  | Rural | 4,588 (21.4) |
| **Relationship status** | Single | 3,422 (15.9) |
|  | Married or civil partnership | 11,900 (55.4) |
|  | Cohabiting | 2,446 (11.4) |
|  | Separated or divorced | 2,060 (9.6) |
|  | Widowed or surviving partner | 1,645 (7.7) |
| **Educational qualifications** | Degree or equivalent qualification | 6,281 (29.3) |
|  | Below degree qualification | 10,866 (50.7) |
|  | No qualification | 4,303 (20.1) |
| **National Statistics Socio-economic Classification (NS-SEC)** | Managerial and professional occupations | 9,387 (43.7) |
|  | Intermediate occupations | 2,384 (11.1) |
|  | Small employers and own account workers | 2,366 (11.0) |
|  | Lower supervisory and technical occupations | 1,738 (8.1) |
|  | Semi-routine occupations | 5,135 (23.9) |
|  | Other | 447 (2.1) |
| **Body Mass Index (BMI)** | Not overweight or obese | 6,502 (33.6) |
|  | Overweight | 7,223 (37.4) |
|  | Obese | 5,602 (29.0) |
| **Self-reported general health** | Very good or good | 15,803 (73.6) |
|  | Fair | 4,010 (18.7) |
|  | Bad or very bad | 1,658 (7.7) |
| **Smoking status** | Current cigarette smoker | 2,546 (11.9) |
|  | Ex-regular cigarette smoker | 6,356 (29.6) |
|  | Never regular cigarette smoker | 12,536 (58.5) |

### Percentage with total hypertension and undiagnosed hypertension by characteristics

Tables 2 and 3 present the percentage of men and women with hypertension and undiagnosed hypertension by characteristics. Among all men, 34.3% (95% confidence interval 33.2-35.4%) had hypertension and 30.7% (29.0-32.4%) of those were undiagnosed. Among all women, 29.7% (28.8-30.7%) had hypertension and 27.6% (26.1-29.1%) of those were undiagnosed.

**Table 2.** Percentage with hypertension and undiagnosed hypertension by characteristics - females

|  | Total hypertension<br>(among all females) |  | Undiagnosed hypertension<br>(among females with hypertension) |  |
| --- | --- | --- | --- | --- |
|  | Weighted N | Percentage<br>(95% CI) | Weighted N | Percentage<br>(95% CI) |
| **Total** | 10890 | 29.7 (28.8 - 30.7) | 3770 | 27.6 (26.1 - 29.1) |
| **Age group** |  |  |  |  |
| 16-24 years | 1350 | 4.0 (2.4 - 5.6) | 60 | 26.1 (8.0 - 44.3) |
| 25-34 years | 1740 | 7.9 (6.3 - 9.5) | 160 | 44.0 (33.3 - 54.6) |
| 35-44 years | 1670 | 13.6 (11.9 - 15.3) | 270 | 34.0 (27.6 - 40.3) |
| 45-54 years | 1880 | 25.9 (23.8 - 27.9) | 570 | 31.0 (27.0 - 35.0) |
| 55-64 years | 1600 | 40.0 (37.8 - 42.3) | 740 | 29.1 (25.8 - 32.4) |
| 65-74 years | 1430 | 57.8 (55.6 - 60.1) | 960 | 26.8 (24.2 - 29.5) |
| 75 years and over | 1220 | 71.2 (68.9 - 73.5) | 1010 | 21.1 (18.6 - 23.5) |
| **Ethnicity** |  |  |  |  |
| White | 9390 | 28.6 (27.3 - 29.8) | 3400 | 22.3 (20.7 - 24.1) |
| Black | 360 | 46.2 (39.9 - 52.6) | 130 | 16.1 (10.7 - 23.6) |
| Asian | 880 | 31.9 (26.8 - 37.4) | 190 | 26.1 (18.8 - 34.9) |
| Other | 260 | 30.8 (23.1 - 39.8) | 50 | 15.3 (8.2 - 26.8) |
| **Region** |  |  |  |  |
| North East | 530 | 31.1 (28.0 - 34.4) | 210 | 22.2 (18.3 - 26.7) |
| North West | 1430 | 31.3 (28.4 - 34.3) | 510 | 21.4 (18.3 - 25.0) |
| Yorkshire and the Humber | 1070 | 29.8 (26.5 - 33.4) | 360 | 19.0 (15.2 - 23.4) |
| East Midlands | 910 | 33.1 (30.1 - 36.2) | 360 | 24.7 (20.8 - 29.0) |
| West Midlands | 1120 | 31.4 (28.1 - 34.8) | 420 | 20.7 (16.8 - 25.1) |
| East of England | 1200 | 28.1 (25.5 - 30.8) | 440 | 21.6 (18.1 - 25.6) |
| London | 1590 | 28.5 (25.2 - 32.1) | 410 | 21.0 (16.6 - 26.3) |
| South East | 1850 | 26.6 (24.4 - 29.0) | 620 | 22.9 (19.7 - 26.4) |
| South West | 1180 | 28.3 (25.4 - 31.4) | 440 | 26.0 (22.1 - 30.4) |
| **Rural-urban classification** |  |  |  |  |
| Urban | 8780 | 29.9 (28.6 - 31.2) | 2940 | 21.6 (19.8 - 23.4) |
| Rural | 2110 | 27.6 (25.4 - 29.8) | 830 | 24.6 (21.6 - 27.8) |
| **Relationship status** |  |  |  |  |
| Single | 2130 | 33.6 (30.2 - 37.3) | 330 | 17.1 (13.5 - 21.4) |
| Married or civil partnership | 5420 | 28.4 (26.8 - 30.1) | 2040 | 24.4 (22.1 - 26.9) |
| Cohabiting | 1400 | 27.3 (24.1 - 30.8) | 250 | 22.3 (17.6 - 27.9) |
| Separated or divorced | 1030 | 29.1 (26.5 - 31.9) | 460 | 21.9 (18.3 - 25.9) |
| Widowed or surviving partner | 910 | 31.2 (27.8 - 34.7) | 700 | 19.7 (16.3 - 23.7) |
| **Educational qualifications** |  |  |  |  |
| Degree or equivalent qualification | 3200 | 23.6 (21.9 - 25.5) | 670 | 26.2 (23.0 - 29.6) |
| Below degree qualification | 5580 | 30.1 (28.6 - 31.6) | 1860 | 22.2 (20.2 - 24.4) |
| No qualification | 2090 | 35.5 (32.8 - 38.3) | 1230 | 18.6 (16.0 - 21.7) |
| **Socio-economic status (NS-SEC)** |  |  |  |  |
| Managerial and professional occupations | 4390 | 25.6 (24.1 - 27.2) | 1290 | 25.0 (22.6 - 27.6) |
| Intermediate occupations | 1340 | 30.6 (27.9 - 33.5) | 550 | 21.2 (17.8 - 25.0) |
| Small employers and own account workers | 1150 | 24.4 (21.7 - 27.4) | 340 | 27.4 (22.7 - 32.5) |
| Lower supervisory and technical occupations | 810 | 31.1 (27.6 - 34.9) | 280 | 21.1 (16.6 - 26.4) |
| Semi-routine occupations | 2810 | 35.5 (33.3 - 37.8) | 1180 | 18.7 (16.4 - 21.3) |
| Other | 360 | 38.2 (30.5 - 46.6) | 120 | 19.2 (11.8 - 29.7) |
| **Body Mass Index (BMI)** |  |  |  |  |
| Not overweight or obese | 4000 | 17.8 (16.3 - 19.3) | 780 | 24.9 (21.7 - 28.4) |
| Overweight | 3090 | 26.7 (24.9 - 28.5) | 1100 | 24.1 (21.4 - 27.0) |
| Obese | 2730 | 43.9 (41.6 - 46.2) | 1390 | 20.5 (18.2 - 22.9) |
| **Self-reported general health** |  |  |  |  |
| Very good or good | 8150 | 25.3 (24.1 - 26.5) | 2260 | 27.5 (25.4 - 29.8) |
| Fair | 1910 | 39.6 (37.0 - 42.4) | 1020 | 14.4 (12.4 - 16.7) |
| Bad or very bad | 830 | 45.3 (41.1 - 49.5) | 500 | 13.9 (11.1 - 17.4) |
| **Cigarette smoking status** |  |  |  |  |
| Current cigarette smoker | 1400 | 29.6 (26.5 - 32.8) | 380 | 22.2 (18.3 - 26.6) |
| Ex-regular cigarette smoker | 2550 | 30.7 (28.7 - 32.8) | 1140 | 20.7 (18.3 - 23.4) |
| Never regular cigarette smoker | 6910 | 29.0 (27.7 - 30.5) | 2260 | 23.3 (21.3 - 25.5) |

All estimates are weighted, and all are age-adjusted except where the predictor is age or where there is no predictor (i.e., total population estimates). Weighted sample sizes are rounded to the nearest 10. Base population for total hypertension is all females; base population for undiagnosed hypertension is females with hypertension.

**Table 3.**
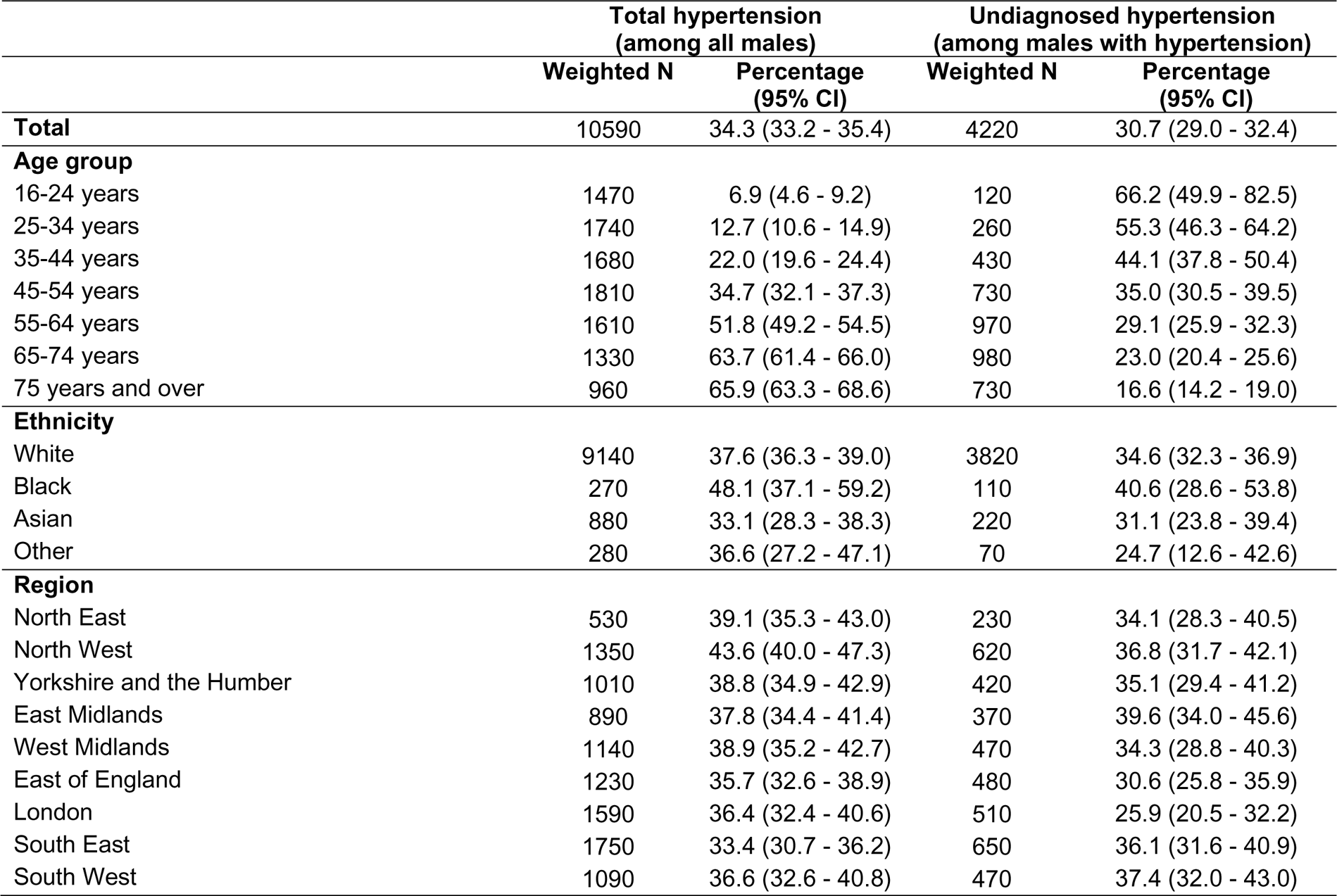
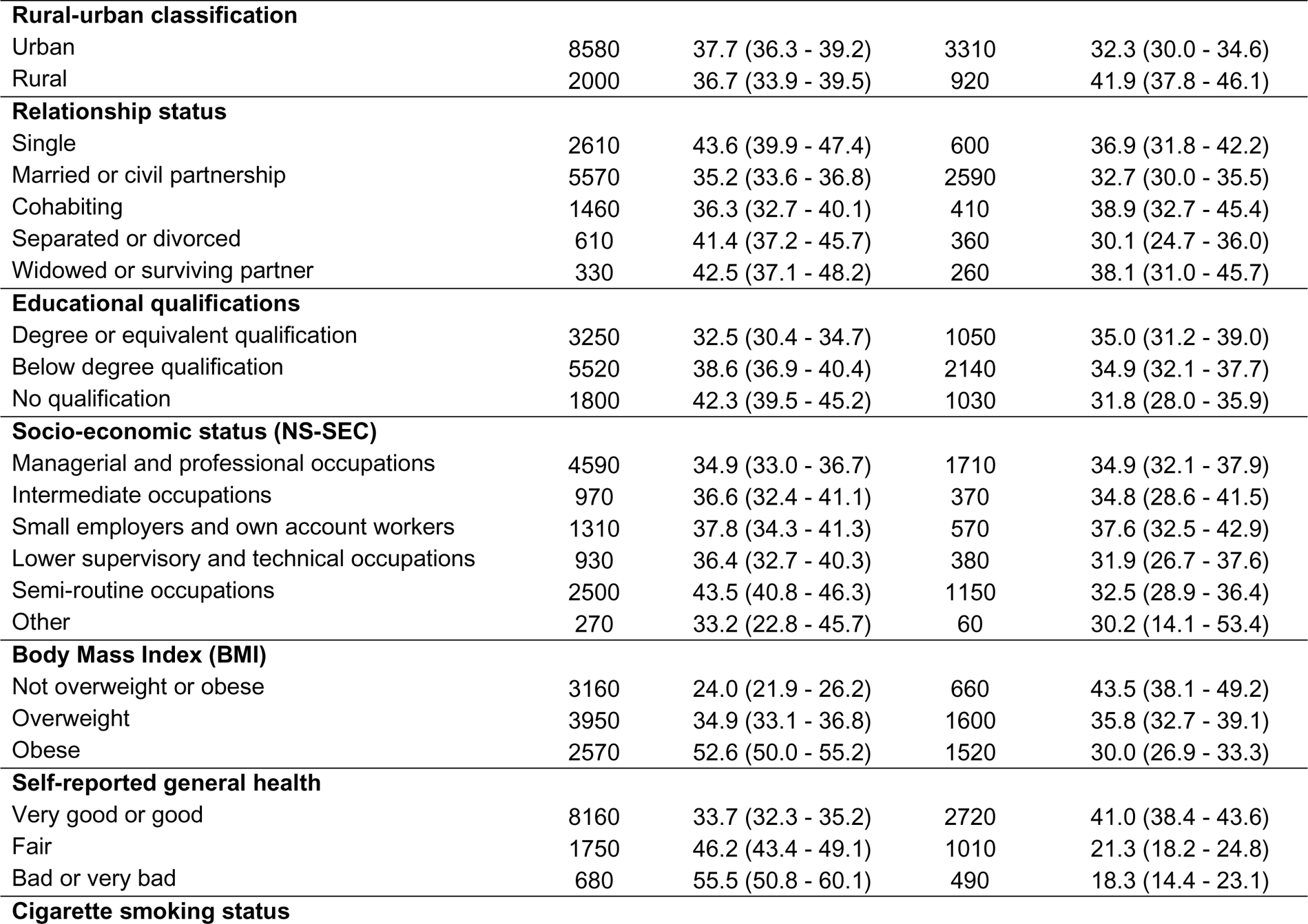
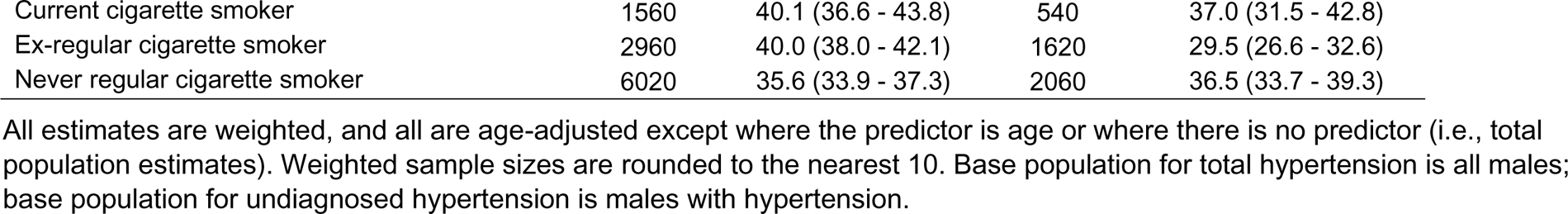
Percentage with hypertension and undiagnosed hypertension by characteristics - males

|  | Total hypertension<br>(among all males) |  | Undiagnosed hypertension<br>(among males with hypertension) |  |
| --- | --- | --- | --- | --- |
|  | Weighted N | Percentage<br>(95% CI) | Weighted N | Percentage<br>(95% CI) |
| **Total** | 10590 | 34.3 (33.2 - 35.4) | 4220 | 30.7 (29.0 - 32.4) |
| **Age group** |  |  |  |  |
| 16-24 years | 1470 | 6.9 (4.6 - 9.2) | 120 | 66.2 (49.9 - 82.5) |
| 25-34 years | 1740 | 12.7 (10.6 - 14.9) | 260 | 55.3 (46.3 - 64.2) |
| 35-44 years | 1680 | 22.0 (19.6 - 24.4) | 430 | 44.1 (37.8 - 50.4) |
| 45-54 years | 1810 | 34.7 (32.1 - 37.3) | 730 | 35.0 (30.5 - 39.5) |
| 55-64 years | 1610 | 51.8 (49.2 - 54.5) | 970 | 29.1 (25.9 - 32.3) |
| 65-74 years | 1330 | 63.7 (61.4 - 66.0) | 980 | 23.0 (20.4 - 25.6) |
| 75 years and over | 960 | 65.9 (63.3 - 68.6) | 730 | 16.6 (14.2 - 19.0) |
| **Ethnicity** |  |  |  |  |
| White | 9140 | 37.6 (36.3 - 39.0) | 3820 | 34.6 (32.3 - 36.9) |
| Black | 270 | 48.1 (37.1 - 59.2) | 110 | 40.6 (28.6 - 53.8) |
| Asian | 880 | 33.1 (28.3 - 38.3) | 220 | 31.1 (23.8 - 39.4) |
| Other | 280 | 36.6 (27.2 - 47.1) | 70 | 24.7 (12.6 - 42.6) |
| **Region** |  |  |  |  |
| North East | 530 | 39.1 (35.3 - 43.0) | 230 | 34.1 (28.3 - 40.5) |
| North West | 1350 | 43.6 (40.0 - 47.3) | 620 | 36.8 (31.7 - 42.1) |
| Yorkshire and the Humber | 1010 | 38.8 (34.9 - 42.9) | 420 | 35.1 (29.4 - 41.2) |
| East Midlands | 890 | 37.8 (34.4 - 41.4) | 370 | 39.6 (34.0 - 45.6) |
| West Midlands | 1140 | 38.9 (35.2 - 42.7) | 470 | 34.3 (28.8 - 40.3) |
| East of England | 1230 | 35.7 (32.6 - 38.9) | 480 | 30.6 (25.8 - 35.9) |
| London | 1590 | 36.4 (32.4 - 40.6) | 510 | 25.9 (20.5 - 32.2) |
| South East | 1750 | 33.4 (30.7 - 36.2) | 650 | 36.1 (31.6 - 40.9) |
| South West | 1090 | 36.6 (32.6 - 40.8) | 470 | 37.4 (32.0 - 43.0) |
| **Rural-urban classification** |  |  |  |  |
| Urban | 8580 | 37.7 (36.3 - 39.2) | 3310 | 32.3 (30.0 - 34.6) |
| Rural | 2000 | 36.7 (33.9 - 39.5) | 920 | 41.9 (37.8 - 46.1) |
| **Relationship status** |  |  |  |  |
| Single | 2610 | 43.6 (39.9 - 47.4) | 600 | 36.9 (31.8 - 42.2) |
| Married or civil partnership | 5570 | 35.2 (33.6 - 36.8) | 2590 | 32.7 (30.0 - 35.5) |
| Cohabiting | 1460 | 36.3 (32.7 - 40.1) | 410 | 38.9 (32.7 - 45.4) |
| Separated or divorced | 610 | 41.4 (37.2 - 45.7) | 360 | 30.1 (24.7 - 36.0) |
| Widowed or surviving partner | 330 | 42.5 (37.1 - 48.2) | 260 | 38.1 (31.0 - 45.7) |
| **Educational qualifications** |  |  |  |  |
| Degree or equivalent qualification | 3250 | 32.5 (30.4 - 34.7) | 1050 | 35.0 (31.2 - 39.0) |
| Below degree qualification | 5520 | 38.6 (36.9 - 40.4) | 2140 | 34.9 (32.1 - 37.7) |
| No qualification | 1800 | 42.3 (39.5 - 45.2) | 1030 | 31.8 (28.0 - 35.9) |
| **Socio-economic status (NS-SEC)** |  |  |  |  |
| Managerial and professional occupations | 4590 | 34.9 (33.0 - 36.7) | 1710 | 34.9 (32.1 - 37.9) |
| Intermediate occupations | 970 | 36.6 (32.4 - 41.1) | 370 | 34.8 (28.6 - 41.5) |
| Small employers and own account workers | 1310 | 37.8 (34.3 - 41.3) | 570 | 37.6 (32.5 - 42.9) |
| Lower supervisory and technical occupations | 930 | 36.4 (32.7 - 40.3) | 380 | 31.9 (26.7 - 37.6) |
| Semi-routine occupations | 2500 | 43.5 (40.8 - 46.3) | 1150 | 32.5 (28.9 - 36.4) |
| Other | 270 | 33.2 (22.8 - 45.7) | 60 | 30.2 (14.1 - 53.4) |
| **Body Mass Index (BMI)** |  |  |  |  |
| Not overweight or obese | 3160 | 24.0 (21.9 - 26.2) | 660 | 43.5 (38.1 - 49.2) |
| Overweight | 3950 | 34.9 (33.1 - 36.8) | 1600 | 35.8 (32.7 - 39.1) |
| Obese | 2570 | 52.6 (50.0 - 55.2) | 1520 | 30.0 (26.9 - 33.3) |
| **Self-reported general health** |  |  |  |  |
| Very good or good | 8160 | 33.7 (32.3 - 35.2) | 2720 | 41.0 (38.4 - 43.6) |
| Fair | 1750 | 46.2 (43.4 - 49.1) | 1010 | 21.3 (18.2 - 24.8) |
| Bad or very bad | 680 | 55.5 (50.8 - 60.1) | 490 | 18.3 (14.4 - 23.1) |
| **Cigarette smoking status** |  |  |  |  |
| Current cigarette smoker | 1560 | 40.1 (36.6 - 43.8) | 540 | 37.0 (31.5 - 42.8) |
| Ex-regular cigarette smoker | 2960 | 40.0 (38.0 - 42.1) | 1620 | 29.5 (26.6 - 32.6) |
| Never regular cigarette smoker | 6020 | 35.6 (33.9 - 37.3) | 2060 | 36.5 (33.7 - 39.3) |

All estimates are weighted, and all are age-adjusted except where the predictor is age or where there is no predictor (i.e., total population estimates). Weighted sample sizes are rounded to the nearest 10. Base population for total hypertension is all males; base population for undiagnosed hypertension is males with hypertension.

Hypertension prevalence increased with age, however, among those with hypertension, younger adults were more likely to be undiagnosed. The groups with the highest proportion undiagnosed were 16-24-year-old men (66.2% (49.9-82.6%)) and 25-34-year-old women (44.0% (33.3-55.6%)). This compared to 16.6% (14.2-19.0%) of men and 21.1% (18.6-23.5%) of women aged 75 and over.

For ethnicity, age-adjusted hypertension prevalence was highest among women who belonged to the Black ethnic group. There were no differences in hypertension prevalence by ethnicity for men, nor in the proportion who were undiagnosed for either men or women.

Men living in the North West of England and women living in the East Midlands had the highest age-adjusted hypertension prevalence, and men and women in the South East had the lowest prevalence. There were no differences in the proportion of women who were undiagnosed by region, but for men the highest proportion undiagnosed was in the East Midlands (39.6% (34.0-45.6%)) and the lowest in London (25.9% (20.5-32.2%) and the East of England (30.6% (25.8-35.9%)). Men living in rural areas were also more likely to be undiagnosed (41.9% (37.8-46.1%)) than those living in urban areas (32.3% (30.0-34.6%)).

Age-adjusted hypertension prevalence was highest among men and women in ‘semi-routine occupations’ and lowest among men and women in ‘managerial and professional occupations’ and women working for ‘small employers and own account workers’. For women, the highest percentage undiagnosed was among ‘small employers and own account workers’ (27.4% (22.7-32.5%)) and ‘managerial and professional occupations’ (25.0% (22.6-27.6%) and the lowest among ‘semi-routine occupations’ (18.7% (16.4-21.3%)), while there were no differences in the proportion undiagnosed by NS-SEC for men.

Men and women with no educational qualifications had the highest age-adjusted hypertension prevalence and those with degree-level or equivalent qualifications had the lowest prevalence. For women, the highest percentage undiagnosed was among those with degree-level or higher qualifications (26.2% (23.0-30.0%)) and the lowest was among those with no qualifications (18.6% (16.0-21.7%)), while there were no differences in the proportion of men undiagnosed by education.

Men and women who were single had the highest age-adjusted hypertension prevalence and those who were married or in a civil partnership had the lowest prevalence. For women, the highest percentage undiagnosed was among those who were married or in a civil partnership (24.4% (22.1-26.9%)) and the lowest was among those who were single (17.1% (13.5-21.4%)), while there were no differences by relationship status for men.

Age-adjusted hypertension prevalence was highest among men and women whose self-reported general health was ‘bad or very bad’ and lowest among those whose health was ‘very good or good’. However, among those with hypertension, adults whose health was ‘very good or good’ were the most likely to be undiagnosed (men 41.0% (38.4-43.6%), women 27.5% (25.4-29.8%)) and those whose health was ‘bad or very bad’ were the least likely to be undiagnosed (men 18.3% (14.4-23.1%), women 13.9% (11.1-17.4%)).

Men and women in the BMI category ‘obese’ had the highest age-adjusted hypertension prevalence and those classified as ‘not overweight or obese’ had the lowest prevalence. However, among those with hypertension, individuals classified as ‘not overweight or obese’ were the most likely to be undiagnosed (men 43.5% (38.1-49.2%), women 24.9% (21.7-28.4%)) and those classified as ‘obese’ were the least likely (men 30.0% (26.9-33.3%), women 20.1% (18.2-22.9%)).

Men who were ex-regular smokers were the most likely to have hypertension and those who had never regularly smoked were the least likely, however, among men with hypertension, those who had never regularly smoked were the most likely to be undiagnosed (36.5% (33.7-39.3%)) and ex-regular smokers were the least likely (29.5% (26.6-32.6%)). There were no differences in prevalence of hypertension or undiagnosed hypertension by smoking status for women.

### Regression models

In fully adjusted models, age, BMI, and self-reported general health were independently associated with the likelihood of men and women with hypertension being undiagnosed after adjusting for sociodemographic characteristics (see Table 4). Compared with those aged 75 years and over, the age groups with the highest odds of being undiagnosed were 16–24-year-old men (odds ratio (OR) 8.5 (3.9-18.7)) and 25–34-year-old women (OR 2.8 (1.7-4.5)). Men and women who were ‘not overweight or obese’ according to their BMI had 1.9 (1.4-2.4) and 1.2 (1.0-1.5) times the odds of being undiagnosed, respectively, compared with those who were classified as ‘obese’. Men and women whose self-reported general health was ‘very good or good’ had 3.5 (2.5-4.8) and 2.2 (1.7-2.9) times the odds of being undiagnosed compared with those whose self-reported general health was ‘bad or very bad’.

**Table 4.** Adjusted odds ratios for undiagnosed hypertension among those with hypertension, by characteristics

|  | Males |  | Females |  |
| --- | --- | --- | --- | --- |
|  | Minimally adjusted model | Fully adjusted model | Minimally adjusted model | Fully adjusted model |
| Age group |  |  |  |  |
| 16-24 years | 9.84 (4.60 - 21.04) | 8.52 (3.89 - 18.67) | 1.32 (0.51 - 3.43) | 1.43 (0.53 - 3.90) |
| 25-34 years | 6.19 (4.13 - 9.28) | 6.10 (3.91 - 9.50) | 2.94 (1.86 - 4.64) | 2.75 (1.66 - 4.54) |
| 35-44 years | 3.96 (2.90 - 5.40) | 4.15 (2.95 - 5.83) | 1.93 (1.40 - 2.65) | 1.70 (1.18 - 2.43) |
| 45-54 years | 2.70 (2.07 - 3.51) | 2.79 (2.09 - 3.73) | 1.68 (1.33 - 2.13) | 1.48 (1.12 - 1.95) |
| 55-64 years | 2.06 (1.63 - 2.60) | 2.14 (1.66 - 2.76) | 1.53 (1.24 - 1.91) | 1.32 (1.04 - 1.69) |
| 65-74 years | 1.50 (1.20 - 1.87) | 1.52 (1.21 - 1.91) | 1.37 (1.12 - 1.68) | 1.27 (1.02 - 1.57) |
| 75 years and over | Reference Group |  |  |  |
| Ethnicity |  |  |  |  |
| Black | 1.25 (0.73 - 2.13) | 1.53 (0.86 - 2.75) | 0.69 (0.42 - 1.11) | 0.80 (0.48 - 1.31) |
| Asian | 0.87 (0.59 - 1.27) | 1.11 (0.74 - 1.65) | 1.25 (0.81 - 1.93) | 1.24 (0.80 - 1.93) |
| Other | 0.68 (0.31 - 1.47) | 0.75 (0.37 - 1.53) | 0.65 (0.32 - 1.32) | 0.67 (0.32 - 1.40) |
| White | Reference Group |  |  |  |
| Region |  |  |  |  |
| North East | 0.77 (0.54 - 1.09) | 0.79 (0.55 - 1.13) | 0.88 (0.64 - 1.21) | 0.87 (0.64 - 1.20) |
| North West | 0.88 (0.64 - 1.21) | 0.92 (0.66 - 1.28) | 0.84 (0.63 - 1.12) | 0.83 (0.61 - 1.11) |
| Yorkshire and the Humber | 0.81 (0.58 - 1.15) | 0.85 (0.60 - 1.20) | 0.72 (0.52 - 1.01) | 0.70 (0.50 - 0.98) |
| West Midlands | 0.80 (0.57 - 1.12) | 0.82 (0.58 - 1.17) | 0.81 (0.59 - 1.12) | 0.81 (0.59 - 1.12) |
| East of England | 0.67 (0.48 - 0.94) | 0.63 (0.45 - 0.88) | 0.85 (0.63 - 1.15) | 0.81 (0.60 - 1.10) |
| London | 0.54 (0.37 - 0.78) | 0.56 (0.39 - 0.83) | 0.85 (0.59 - 1.21) | 0.82 (0.57 - 1.18) |
| South East | 0.86 (0.64 - 1.17) | 0.86 (0.63 - 1.17) | 0.92 (0.70 - 1.22) | 0.88 (0.66 - 1.17) |
| South West | 0.89 (0.64 - 1.25) | 0.83 (0.59 - 1.17) | 1.07 (0.79 - 1.43) | 1.01 (0.75 - 1.36) |
| East Midlands | Reference Group |  |  |  |
| Rural-urban classification |  |  |  |  |
| Rural | 1.50 (1.24 - 1.81) | 1.47 (1.21 - 1.79) | 1.17 (0.98 - 1.39) | 1.07 (0.89 - 1.29) |
| Urban | Reference Group |  |  |  |
| Relationship status |  |  |  |  |
| Married or civil partnership | 0.85 (0.65 - 1.09) | 0.81 (0.62 - 1.05) | 1.61 (1.17 - 2.22) | 1.43 (1.04 - 1.98) |
| Cohabiting | 1.09 (0.76 - 1.56) | 1.03 (0.71 - 1.49) | 1.45 (0.97 - 2.16) | 1.34 (0.90 - 2.00) |
| Separated or divorced | 0.75 (0.53 - 1.05) | 0.74 (0.52 - 1.04) | 1.41 (0.98 - 2.03) | 1.37 (0.95 - 1.97) |
| Widowed or surviving partner | 1.11 (0.76 - 1.61) | 1.08 (0.74 - 1.59) | 1.19 (0.82 - 1.73) | 1.17 (0.80 - 1.69) |
| Single | Reference Group |  |  |  |
| Educational qualifications |  |  |  |  |
| Degree or equivalent qualification | 1.15 (0.90 - 1.47) | 1.13 (0.86 - 1.48) | 1.56 (1.21 - 2.02) | 1.37 (1.05 - 1.79) |
| Below degree qualification | 1.14 (0.94 - 1.40) | 1.10 (0.89 - 1.36) | 1.26 (1.03 - 1.53) | 1.18 (0.97 - 1.44) |
| No qualification | Reference Group |  |  |  |
| Socio-economic status (NS-SEC) |  |  |  |  |
| Managerial and professional occupations | 1.13 (0.92 - 1.39) | 1.12 (0.89 - 1.41) | 1.42 (1.17 - 1.72) | 1.23 (1.00 - 1.52) |
| Intermediate occupations | 1.12 (0.81 - 1.55) | 1.11 (0.78 - 1.57) | 1.14 (0.89 - 1.46) | 1.09 (0.84 - 1.41) |
| Small employers and own account workers | 1.26 (0.97 - 1.66) | 1.28 (0.97 - 1.68) | 1.62 (1.24 - 2.13) | 1.45 (1.09 - 1.92) |
| Lower supervisory and technical occupations | 0.99 (0.74 - 1.33) | 1.01 (0.75 - 1.36) | 1.15 (0.83 - 1.58) | 1.08 (0.78 - 1.51) |
| Other | 0.96 (0.38 - 2.45) | 1.05 (0.46 - 2.41) | 1.06 (0.59 - 1.88) | 1.16 (0.66 - 2.02) |
| Semi-routine occupations | Reference Group |  |  |  |
| Body Mass Index (BMI) |  |  |  |  |
| Not overweight or obese | 1.78 (1.38 - 2.29) | 1.86 (1.44 - 2.40) | 1.28 (1.04 - 1.58) | 1.24 (1.00 - 1.53) |
| Overweight | 1.30 (1.08 - 1.56) | 1.30 (1.08 - 1.58) | 1.23 (1.02 - 1.47) | 1.20 (1.00 - 1.44) |
| Obese | Reference Group |  |  |  |
| Self-reported general health |  |  |  |  |
| Very good or good | 3.11 (2.30 - 4.20) | 3.46 (2.52 - 4.75) | 2.32 (1.79 - 3.02) | 2.21 (1.70 - 2.87) |
| Fair | 1.21 (0.86 - 1.70) | 1.29 (0.91 - 1.82) | 1.02 (0.76 - 1.38) | 1.00 (0.75 - 1.35) |
| Bad or very bad | Reference Group |  |  |  |
| Cigarette smoking status |  |  |  |  |
| Ex-regular cigarette smoker | 0.72 (0.55 - 0.94) | 0.71 (0.54 - 0.93) | 0.93 (0.70 - 1.23) | 0.84 (0.63 - 1.11) |
| Never regular cigarette smoker | 0.98 (0.75 - 1.27) | 0.95 (0.73 - 1.24) | 1.07 (0.82 - 1.40) | 0.95 (0.73 - 1.24) |
| Current cigarette smoker | Reference Group |  |  |  |

Minimally adjusted models were adjusted for age group and stratified by sex. Fully adjusted models were adjusted for age group, ethnicity, region, rural-urban classification, relationship status, highest educational qualification, and NS-SEC, and stratified by sex

Among men with hypertension, other characteristics that were independently associated with the likelihood of being undiagnosed were rural-urban classification (rural OR 1.5 (1.2-1.8) compared with urban), region (London OR 0.6 (0.4-0.8), East of England OR 0.6 (0.5-0.9) compared with East Midlands), and smoking status (ex-regular smokers OR 0.7 (0.5-0.9) compared with current smokers). Among women with hypertension, other characteristics that were independently associated with the likelihood of being undiagnosed were NS-SEC (‘small employers and own account workers’ OR 1.5 (1.1-1.9), ‘managerial and professional occupations’ OR 1.2 (1.0-1.5) compared with ‘semi-routine occupations’), educational qualifications (degree-level qualification or equivalent OR 1.4 (1.1-1.8) compared with no qualification), and relationship status (married or in a civil partnership OR 1.4 (1.0-2.0) compared with single).

## Discussion

This study found that many of the groups who were less likely to have hypertension, such as younger adults and those with a lower BMI, were more likely to be undiagnosed if they did have hypertension. Our finding that 30.7% of men and 27.6% of women with hypertension were undiagnosed aligns with previous estimates in England, ^[10]^ and provides further evidence for the substantial burden of undiagnosed hypertension, which is likely to have further increased since the COVID-19 pandemic. ^[12–13]^ Our findings suggest that current approaches to hypertension detection are targeting those most at risk, but also indicate that considerable numbers of young and otherwise healthy individuals have undiagnosed hypertension. Given the high lifetime risk of hypertension and its strong links with morbidity and mortality, ^[1–2]^ our results suggest a need for greater awareness of the potential for undiagnosed hypertension among all groups, including those considered “low risk”. This will be especially important if the government is to achieve its ambition of diagnosing 80% of hypertension cases in England by 2029. ^[11]^

Individuals with known hypertension risk factors (such as older adults and those in poor health) are likely to have more frequent contact with healthcare services, providing greater opportunity for blood pressure measurement. Those with known risk factors may also be more likely to have their blood pressure opportunistically measured during routine healthcare appointments. In our analysis, men and younger adults were more likely to be undiagnosed than women and older adults, which aligns with previous research. ^[17–18]^ In 2019, 69% of men in England reported having consulted with a GP in the previous 12 months, compared with 82% of women, and sex differences were greater among younger adults. ^[5]^ Women are more likely to have routine healthcare appointments where blood pressure is measured, for example during oral contraceptive check-ups or during pregnancy or post-natal healthcare checks. However, research has shown that men have fewer healthcare consultations than women even after accounting for reproductive-related contacts.^[19]^ This suggests that relying solely on contact with healthcare services for hypertension detection is likely to systematically overlook certain groups of the population, including young men.

Better self-reported general health and a lower BMI were associated with an increased likelihood of hypertension being undiagnosed. This is likely due to the relationships between general health, BMI and a variety of long-term health conditions and may therefore be related to frequency of contact with healthcare services or the likelihood of opportunistic blood pressure measurement taking place during those contacts. However, individuals with a diagnosis of hypertension may factor this into self-reports of their general health and reverse causality or a bidirectional relationship is likely. That is to say that individuals who are otherwise in good health may be less likely to have their hypertension diagnosed, but individuals who do not know they have hypertension may also be more likely to self-report their health as good.

Men who lived in London and the East of England, and those who lived in urban areas, were less likely to be undiagnosed than those living in other regions or in rural areas. A previous study found that rates of hypertension diagnosis were lower among people living in rural areas in low- and middle-income countries but not in high-income countries, although the analysis was not disaggregated by sex. ^[17]^ Our findings suggest that convenience of access to healthcare may be a significant factor for hypertension diagnosis among men.

Among women, additional factors that were associated with a higher likelihood of hypertension being undiagnosed were being more highly educated, being married or in a civil partnership, and belonging to the NS-SEC classes ‘managerial and professional occupations’ or ‘small employers or own account workers’. Individuals who are more highly educated and those who belong to higher socioeconomic classes have better health on average than those who are less educated or who belong to lower socioeconomic classes, ^[20–21]^ and may therefore have reduced opportunity for blood pressure monitoring as part of routine healthcare. However, in our analysis, education and NS-SEC were only associated with undiagnosed hypertension for women. Analysis of primary care data in the UK found a deprivation gradient in healthcare use among women whereby those in more deprived areas had higher rates of GP consultations than women in less deprived areas, but no difference was found for men, ^[19]^ which may explain our findings. Women with higher levels of education also have fewer children on average, ^[22]^ and differences in reproductive-related healthcare contacts may add to the deprivation effect for women. The difference in undiagnosed hypertension by marital status among women is unexpected and warrants further investigation.

The Health Survey for England (HSE) provided a unique opportunity to explore inequalities in undiagnosed hypertension. The rigorous survey design and weighting methods enabled us to produce estimates of undiagnosed hypertension for a representative sample of adults in England. The survey benefits from stringent blood pressure protocols designed to optimise the accuracy of readings. Nonetheless, our study had limitations. Firstly, despite efforts to boost sample size by pooling data from multiple years, limitations arose from small numbers with hypertension in some predictor groups. This was a particular issue for ethnicity, which led to wide confidence intervals for the “Black”, “Asian”, and “Other” ethnic groups, especially for men. This may explain why we found little evidence for differences in hypertension prevalence by ethnicity for men, despite previous literature demonstrating higher prevalence among Black men as well as Black women. ^[15]^ Secondly, our analysis used self-reports of hypertension which will not fully align with clinical diagnoses. Research on self-reports of hypertension offer mixed evidence for their accuracy, ^[23–24]^ but some suggest underestimation compared to clinical records, which would imply overestimation of undiagnosed hypertension in our analysis. However, this would only impact the interpretation of our findings if the level of disagreement varied across groups. Other studies use self-reports of anti-hypertensive medication instead of self-reports of diagnosis, ^[5, 25]^ however this excludes individuals with hypertension controlled by lifestyle interventions. Blood pressure measurements taken at a single point in time are not equivalent to a diagnosis of hypertension, which requires 24-hour ambulatory or at-home blood pressure monitoring. ^[9]^ Because the HSE cannot show individual variability in blood pressure over time, there is potential for low reproducibility of measurements and over-estimation of undiagnosed hypertension. Again, this would only impact interpretation of our findings if conversion of single-point-in-time measurements to clinical diagnoses differed between groups.

In England, blood pressure checks are offered every five years to adults aged 40-75 under the NHS Health Check, and 1 in 34 Health Check attendees are diagnosed with hypertension, but the programme had an uptake of just 53% between 2012 and 2017 and this further reduced to 40% in 2021/22. ^[26–27]^ There is substantial regional disparity in uptake, ranging from 25% to 85%, as well as differences by demographic factors such as sex (higher uptake among females) and age (higher uptake among older adults). Since 2021, free blood pressure checks for adults over the age of 40 have also been available at community pharmacies. Despite these programmes, our findings demonstrate that undiagnosed hypertension remains prevalent. Early identification and intervention are associated with better health and economic outcomes, ^[2–3]^ and there may be substantial benefit in ensuring accessibility of hypertension screening for all groups, including those who are not routinely in contact with healthcare services. A 2015 systematic review emphasised the potential of community-based, non-physician or self-screening methods to improve detection of hypertension, ^[28]^ however, a recent Cochrane review highlighted the need for additional high-quality evidence on the effectiveness of different screening strategies in reducing hypertension-related morbidity and mortality. ^[29]^

This study builds on the evidence for the substantial burden of undiagnosed hypertension in England and provides evidence that some of the groups who are less likely to have hypertension (such as younger adults and those with a lower BMI) are the most likely to be undiagnosed if they do have hypertension. Given the high lifetime risk of hypertension, its strong associations with morbidity and mortality, and the health and economic benefits of early identification and intervention, ^[1–4]^ our findings suggest a need for greater awareness of the risks of undiagnosed hypertension, including among those typically considered “low risk”.

## Data Availability

Data from the Health Survey for England is available for researchers to access via the UK Data Service.

## Contribution of authors

KF, VN and CS conceived the idea for the study. Study design was led by KF and EC with input from CS and VN. EC was responsible for curating and cleaning the data, and analysis was carried out by EC, EM, KF, and NCC. KF and CS had leadership responsibility and supervised the project. Methodological advice was provided by VN, PP and IW. AH, VR and AB provided topic expertise and contributed to validation of methods and findings. KF, EC and EM wrote the first draft of the publication. All authors reviewed and commented on the publication and approved the final version. KF, EC, EM and CS had access to the data and were responsible for the decision to submit for publication.

## Acknowledgements

AB is chief investigator of the NIHR-funded STIMULATE-ICP programme [NIHR COV-LT2-0043]. The funders had no role in study design, data collection and analysis, decision to publish, or preparation of the manuscript.

## References

1. B. Zhou, P. Perel, G. A. Mensah and M. Ezzati. Global epidemiology, health burden and effective interventions for elevated blood pressure and hypertension. Nature Reviews Cardiology 2021, 18(11), 785–802. doi: 10.1038/s41569-021-00559-8

2. NCD Risk Factor Collaboration. Worldwide trends in hypertension prevalence and progress in treatment and control from 1990 to 2019: a pooled analysis of 1201 population-representative studies with 104 million participants. The Lancet 2021, 398(10304), 957–80. doi: 10.1016/S0140-6736(21)01330-1

3. World Health Organization. A global brief on hypertension: Silent killer, global public health crisis. World Health Organization, 2013. Geneva.

4. GBD 2019 Risk Factors Collaborators. Global burden of 87 risk factors in 204 countries and territories, 1990-2019: a systematic analysis for the Global Burden of Disease Study 2019. The Lancet 2020, 396(10258), 1223–1249.

5. NHS Digital. Health Survey for England 2019 (National Statistics). NHS Digital 2019, London. https://digital.nhs.uk/data-and-information/publications/statistical/health-survey-for-england/2019

6. Public Health England. Health matters: preventing cardiovascular disease. 2019. [Online]. https://www.gov.uk/government/publications/health-matters-preventing-cardiovascular-disease/health-matters-preventing-cardiovascular-disease [Accessed May 2023].

7. D. Ettehad, C. A. Emdin, A. Kiran, S. G. Anderson, T. Callender, J. Emberson, J. Chalmers, A. Rodgers and K. Rahimi. Blood pressure lowering for prevention of cardiovascular disease and death: a systematic review and meta-analysis. The Lancet 2016, 387(10022), 957–967. doi: 10.1016/S0140-6736(15)01225-8.

8. M. Brunström and B. Carlberg. Association of Blood Pressure Lowering With Mortality and Cardiovascular Disease Across Blood Pressure Levels. JAMA Internal Medicine 2018, 178(1), 28–36. doi: 10.1001/jamainternmed.2017.6015

9. National Institute for Health and Care Excellence. Hypertension in adults: diagnosis and management. Nice guideline NG136. 2019. [Online]. https://www.nice.org.uk/guidance/ng136 [Accessed May 2023].

10. S. Scholes, A. Conolly and J. S. Mindell. Income-based inequalities in hypertension and undiagnosed hypertension: analysis of Health Survey for England data. Journal of Hypertension, 2020, 38(5), 912–924. doi: 10.1097/HJH.0000000000002350

11. Public Health England and NHS England. Ambitions set to address major causes of cardiovascular disease. 2019. [Online]. https://www.gov.uk/government/news/ambitions-set-to-address-major-causes-of-cardiovascular-disease [Accessed May 2023]

12. CVDPREVENT. CVDPREVENT Third annual report. Office for Health Improvement and Disparities and the NHS Benchmarking Network, 2022, London.

13. C. E. Dale, R. Takhar, R. Carragher, M. Katsoulis, F. Torabi, S. Duffield and et al. The impact of the COVID-19 pandemic on cardiovascular disease prevention and management. Nature Medicine 2023, 29(1), 219–225. doi: 10.1038/s41591-022-02158-7

14. B. Leng, Y. Jin, G. Li, L. Chen and N. Jin. Socioeconomic status and hypertension: A meta-analysis. Journal of Hypertension 2015, 33(2), 221–229. DOI: 10.1097/HJH.0000000000000428

15. NHS Digital. Health Survey England Additional Analyses, Ethnicity and Health, 2011-2019 Experimental statistics. NHS Digital 2022, London. https://digital.nhs.uk/data-and-information/publications/statistical/health-survey-england-additional-analyses/ethnicity-and-health-2011-2019-experimental-statistics

16. B. Williams, G. Mancia, W. Spiering, E. A. Rosei, M. Azizi, M. Burnier, D. L. Clement and et al. 2018 ESC/ESH Guidelines for the management of arterial hypertension: The Task Force for the management of arterial hypertension of the European Society of Cardiology (ESC) and the European Society of Hypertension (ESH). European Heart Journal 2018, 39(33), 3021–3104. doi: 10.1093/eurheartj/ehy339

17. C. K. Chow, K. K. Teo and S. Rangarajan. Prevalence, Awareness, Treatment, and Control of Hypertension in Rural and Urban Communities in High-, Middle-, and Low-Income Countries. JAMA 2013, 310(9), 959–968. doi: 10.1093/gerona/glad155

18. A. I. Uhernik, V. Kralj, P. Čukelj, I. Brkić-Biloš, M. Erceg, T. Benjak and R. Stevanović. Undiagnosed hypertension in Croatia. Croatian Medical Journal 2023, 64(1), 4–12. doi: 10.3325/cmj.2023.64.4

19. Y. Wang, K. Hunt, I. Nazareth, N. Freemantle and I. Petersen. Do men consult less than women? An analysis of routinely collected UK general practice data. BMJ Open 2013, 3, e003320. doi: 10.1136/bmjopen-2013-003320

20. Office for National Statistics. An overview of lifestyles and wider characteristics linked to Healthy Life Expectancy in England: June 2017. 2017. [Online]. https://www.ons.gov.uk/peoplepopulationandcommunity/healthandsocialcare/healthinequalities/articles/healthrelatedlifestylesandwidercharacteristicsofpeoplelivinginareaswiththehighestorlowesthealthylife/june2017 [Accessed 11 May 2023].

21. J. P. Mackenbach, I. Stirbu, A.-J. R. Roskam, M. M. Schaap, G. Menvielle, M. Leinsalu and A. E. Kunst. Socioeconomic Inequalities in Health in 22 European Countries. N Engl J Med 2008, 358(23), 2468–2481. doi: 10.1056/NEJMsa0707519

22. A. Berrington, J. Stone and E. Beaujouan. Educational differences in childbearing widen in Britain. 2015. [Online]. http://www.cpc.ac.uk/docs/BP29_Educational_differences_in_childbearing_widen-in_Britain.pdf [Accessed 11 May 2023].

23. V. S. Gonçalves, K. R. Andrade, K. M. Carvalho, M. T. Silva, M. G. Pereira and T. F. Galvao. Accuracy of self-reported hypertension: a systematic review and meta-analysis. Journal of Hypertension 2018, 36(5), 970–978. doi: 10.1097/HJH.0000000000001648

24. Y. Okura, L. H. Urban, D. W. Mahoney, S. J. Jacobsen and R. J. Rodeheffer. Agreement between self-report questionnaires and medical record data was substantial for diabetes, hypertension, myocardial infarction and stroke but not for heart failure. Journal of Clinical Epidemiology 2004, 57(10), 1096–1103. doi: 10.1016/j.jclinepi.2004.04.005

25. M. T. Crim, S. S. Yoon, E. Ortiz, H. K. Wall, S. Schober, C. Gillespie, P. Sorlie, N. Keenan, D. Labarthe and Y. Hong. National surveillance definitions for hypertension prevalence and control among adults. Circulation Cardiovascular Quality and Outcomes 2012, 5(3), 343–351. doi: 10.1161/CIRCOUTCOMES.111.963439

26. R. Patel, S. Barnard, K. Thompson, C. Lagord, E. Clegg, R. Worrall, T. Evans, S. Carter, J. Flowers, D. Roberts, M. Nuttall, N. J. Samani, J. Robson, M. Kearney, J. Deanfield and J. Waterall. Evaluation of the uptake and delivery of the NHS Health Check programme in England, using primary care data from 9.5 million people: a cross-sectional study. BMJ Open 2020, 10, e042963. doi: 10.1136/bmjopen-2020-042963

27. Office for Health Improvement and Disparities. NHS Health Check. 2023. [Online]. https://fingertips.phe.org.uk/profile/nhs-health-check-detailed/data#page/1 [Accessed May 2023].

28. S. Fleming, H. Atherton, D. McCartney, J. Hodgkinson, S. Greenfield, F. D. R. Hobbs, J. Mant, R. J. McManus, M. Thompson, A. Ward and C. Heneghan. Self-Screening and Non-Physician Screening for Hypertension in Communities: A Systematic Review. American Journal of Hypertension 2015, 28(11), 1316–1324. doi: 10.1093/ajh/hpv029

29. B.-M. Schmidt, S. Durao, I. Toews, C. M. Bavuma, A. Hohlfeld, E. Nury, J. J. Meerpohl and T. Kredo. Screening strategies for hypertension. Cochrane Database of Systematic Reviews 2020, 5. https://doi.org/10.1002/14651858.CD013212.pub2

