## Supplementary Materials for "Sociodemographic and health-related differences in undiagnosed hypertension in the Health Survey for England 2015-2019"

**Figure S1. Flow chart of sample selection**

Excluded:

Under 16 n=13,922

Pregnant n=207

No valid blood pressure measurement n=18,728

13 participants with invalid or missing data on questions about doctor-diagnosed high blood pressure

54,346

survey respondents

2015 to 2019

21,489 participants eligible for inclusion in analysis

‘Adults with hypertension’ base population for analysis

7,997 participants

‘All adults’ base population for analysis

21,476 participants

21,476 participants included in sample

**Table S1. Characteristics included in analysis**

| **Characteristic** | **Levels** | **Details** |
| --- | --- | --- |
| Sex | Male  Female | Self-reported binary variable |
| Age group | 16-24 years  25-34 years  35-44 years  45-54 years  55-64 years  75+ years | Derived from self-reported age at last birthday |
| Ethnicity | White  Black  Asian  Other | Derived from self-reported ethnicity selected from 17 specified ethnic groups |
| Region | North East  North West  Yorkshire and the Humber  East Midlands  West Midlands  East of England  London  South East  South West | Defined by household address |
| Urban-rural classification | Urban  Rural | Derived variable on rurality of dwelling. Rural areas include town/ fringe/village, hamlet and isolated dwellings. |
| Relationship status | Single  Married or civil partnership  Cohabiting  Separated or divorced  Widowed or surviving partner | Derived from responses to an interview question about relationships within the household. |
| Highest educational qualification | Degree or equivalent qualification  Below degree qualification  No qualification | Derived from self-reported qualifications from a list of 29 options |
| NS-SEC | Managerial and professional occupations  Intermediate occupations  Small employers and own account workers  Lower supervisory and technical occupations  Semi-routine occupations  Other | Derived from responses to questions about employment status and working patterns |
| Body Mass Index | Not overweight or obese  Overweight  Obese | Calculated from height and weight measurements taken by the interviewer, or estimated weight if participants were believed to be above 130kg. BMI <25 not overweight or obese; 25-29 overweight; ≥30 obese. |
| Self-reported general health | Very good or good  Fair  Bad or very bad | Derived from self-assessment of general health on a scale from 1 (very good) to 5 (very bad) |
| Smoking status | Current cigarette smoker  Ex-regular cigarette smoker  Never regular cigarette smoker | Derived from responses to several questions about current and past smoking habits |

NS-SEC - National Statistics Socio-Economic Classification
